# Impact of mHealth interventions on antenatal and postnatal care utilization in low and middle-income countries: A Systematic Review and Meta-Analysis

**DOI:** 10.1101/2020.12.22.20248713

**Authors:** Ravi Kant, Poonam Yadav, Surekha Kishore, Shruti Barnwal, Rajesh Kumar, Meenakshi Khapre

**Author notes:** **Corresponding Author Poonam Yadav**, Ph.D. Scholar, College of Nursing, All India Institute of Medical Sciences, Rishikesh, Uttrakhand, India. Prior publication: -Nil Support: -Nil Conflicts of interest: -Nil Permissions: -NA.

## Abstract

**Objective:** To evaluate the effectiveness of mHealth interventions on antenatal and postnatal care utilization in low and middle-income countries.

**Design:** Systematic review and meta-analysis

**Setting:** Studies from low and middle-income countries were included for analysis.

**Participants:** We searched the literature through major electronic databases such as PubMed, MEDLINE, Embase, Cochrane, Scopus, CINAHL, Clinical key, Google Scholar, Ovid databases with selected keywords, and explored the reference list of articles. Meta-analysis was performed in RevMan 5.4 software; p-value < 0.05 was considered statistically significant. The effect of variables was measured in the Odds ratio with a fixed-effect model. Six published interventional studies were selected as per the eligibility and PICO framed for the systematic review and meta-analysis. Search restricted to articles in the English language, online published, and preprint articles till September 2020.

**Primary and secondary outcome measures:** We evaluated the effectiveness of mHealth intervention on antenatal care utilization, including four antenatal check-ups, iron-folic acid supplementation, two tetanus toxoid immunizations, and postnatal care utilization, which includes postnatal check-ups of delivered mothers.

**Results:** Results have been presented in the form of a forest plot. Findings of this meta-analysis depicted the significant increase in four or more antenatal care attendance (OR=1.89, 95% CI-1.49-2.19), TT immunization (OR=1.63 (95% CI-1.17-2.27), compliance to iron supplementation (OR=1.88, 95% CI-1.18-3.00) and postnatal care attendance (OR=2.54 (95% CI-2.15-2.99) among those pregnant mothers who received mHealth intervention compared to control group.

**Conclusion:** This meta-analysis concluded that m-health has the potential to increase the utilization of full antenatal care and postnatal care compared to standard care, although the level of evidence is moderate.

**Trial registration:** CRD42020204618, PROSPERO, International prospective register of systematic reviews

**Article Summary:** *Strengths and limitations of this study:*

- This meta-analysis creates an evidence for the effectiveness of mHealth with pooled data of interventional studies with limited sample sizes.
- Technology is changing, but even with limited support like SMS, there was an improvement in antenatal and postnatal service utilization.
- Sensitivity analysis identified possible reasons for heterogeneity among studies.
- Studies included from LMICs so results can be generalized for the respective population.
- mHealth as an intervention is a broad term that created heterogeneity also.

## INTRODUCTION

Pregnant women have the right to attain antenatal care throughout the pregnancy for the well-being of the fetus and themselves to promote a positive pregnancy outcome. ^[1]^ Full antenatal care is a term used to deliver appropriate antenatal care to the pregnant mothers, inclusive of four or more antenatal check-ups, and at least one tetanus toxoid injection with consumption of minimum 100 iron-folic acid tablets.^[2]^ A proper antenatal and postnatal check-up provides necessary care to the mother and baby, which helps in timely identification, management, and referral to appropriate facilities to promote a healthy baby’s delivery to a healthy mother to reduce the burden of maternal and neonatal mortality. ^[3]^ According to WHO, nearly 808 women died every day due to preventable causes of pregnancy and childbirth such as hemorrhage, hypertension, infections, and indirect causes, due to the interaction between pre-existing medical conditions and pregnancy and 94% of them from low middle-income countries. ^[4]^ A systematic analysis done by the UN Maternal Mortality Estimation Inter-Agency Group, based on MMR’s sustainable development goal, estimated global projection of MMR for 2030, nearly 161 deaths per 100000 live births. It is estimated as 357 deaths per 100000 live births in Sub-Saharan Africa, 115 deaths per 100000 live births in Southern Asia, 72 deaths per 100000 live births in Southeastern Asia, and 43 deaths per 100000 live births in Northern Africa. ^[5]^ It is also observed that a woman in low-income countries has120 times higher risk of mortality due to pregnancy and childbirth-related causes compared to higher-income countries. ^[4, 6]^ Probable reasons for such bad pregnancy-related outcomes could be administrative, logistic, and technical failures along with insufficient financial support and skilled health personnel, poor accessibility, and non-adherence to maternal and child health services. ^[7]^ The emergence of mobile technology in health facilitates the opportunity to reach the concerned individual and modify their attitude and behavior for a specific health issue. ^[8]^ It can also be an effective tool in sensitizing the population and imparting prenatal education as an engagement tool. ^[9]^ Mobile technologies have effectively created a place through robust communication engagement in treating the disease condition that requires treatment adherence and follow-ups. ^[10]^ It also has the potential to improve perinatal health outcomes by enhancing the acceptability and accessibility of existing maternal and child health services. ^[11]^ We have reviewed relevant studies that have evaluated mobile technology’s effectiveness in promoting maternal and child health by measuring maternal satisfaction, their attitude and behavior towards available maternal and child health services, their antenatal and postnatal attendance, and maternal and perinatal health as well. ^[12-17]^ However, it remains inconclusive due to the limited sample size; hence, it is required to integrate and analyze these studies’ findings with a systematic review of all available relevant literature and meta-analysis. This study evaluated mHealth interventions’ effectiveness in antenatal and postnatal care utilization in low and middle-income countries.

### Aim

To evaluate the effectiveness of mHealth interventions on antenatal and postnatal care utilization in low and middle-income countries.

### Objectives

1. To assess the effectiveness of mHealth interventions on antenatal visits of pregnant women.
2. To assess the effectiveness of mHealth interventions on tetanus toxoid immunization of pregnant women
3. To assess the effectiveness of mHealth interventions on compliance of pregnant women to iron supplementation.
4. To assess the effectiveness of mHealth interventions on postnatal visits of delivered mothers.

## METHODS

### Data Sources and Search strategy

We followed the Preferred Reporting Items for Systematic Review and Meta-Analysis (PRISMA) 2009 guidelines for the systematic review. [Additional file]^[18]^ We reviewed literature from a broad range of databases-PubMed, MEDLINE, Embase, Cochrane, Scopus, CINAHL, Clinical key, Google Scholar, Ovid databases. Search restricted to articles in the English language, online published, and preprint articles till September 2020. The search strategy used the keywords as “pregnant women”, “antenatal attendance”, “ANC visit”, “pregnancy”, “maternal health”, “pregnan*”, “Prenatal Care”, “antenatal mothers”, “postnatal mothers”, “postnatal care”, “tetanus toxoid”, “iron supplementation”, “compliance to iron therapy” AND “mHealth”, “mobile health”, “mobile intervention”, “text message”, “telemedicine”, “mobile technology” AND “LMICs” “resource-limited countries”.

### PICO framework for eligibility

#### Participants

Inclusion criteria: - Pregnant mothers aged 18-49 years, in antenatal, intranatal, and postnatal periods and attended ANC visit in institutional or community setting (PHCs, CHCs, and SCs) Exclusion- Adolescent females

#### Intervention

mHealth -Any intervention designed with a mobile phone or smartphone to support pregnant women and their neonates’ health. It includes text messages, phone calls, and reminders through the app to increase antenatal care of pregnant and postnatal mothers. We excluded the studies which used mobile technology for any other communication purpose or specific disease.

#### Comparator

In the control group, we included conventional or routine services or non-exposure to intervention.

#### Outcome

Antenatal care utilization includes four antenatal check-ups, iron-folic acid supplementation, two tetanus toxoid immunization.

Postnatal care utilization includes postnatal check-ups of delivered mothers.

#### Setting

Inclusion: - Studies conducted in a population of low middle-income countries.

Exclusion: - Studies conducted in high-income countries

#### Time frame

We included studies conducted in the time frame from 2008 to 2020.

#### Study designs

Randomized controlled trials, cluster-randomized controlled trials, quasi-randomized trials, case-control, and cohort studies.

#### Data extraction

After systematic searching of literature, titles and abstracts were screened by two reviewers independently according to inclusion criteria with the help of Rayyan (https://rayyan.qcri.org), a free web-based software.^[19]^ The third reviewer has resolved any discrepancies related to the eligibility of studies. After screening articles, eligible studies were extracted into a data extraction file and imported to RevMan software for meta-analysis. After analysis, data have been presented in the form of forest plots separately for each outcome.

#### Evaluation for the risk of bias

The risk of bias has been evaluated through the Cochrane Risk of Bias Assessment Tool. It is used to assess Random sequence generation (selection bias), Allocation concealment (selection bias), Blinding of participants and personnel (performance bias), Blinding of outcome assessment (detection bias), Incomplete outcome data (attrition bias), Selective reporting (reporting bias) and Other bias through Review Manager software 5.4 version and have been displayed along with forest plot. ^[20]^ Relevant file has been imported from RevMan software to “Summary of Findings” table GRADE Profiler to create a “Summary of Findings” table. ^[21]^ A summary of the intervention effect and a measure of the quality of evidence was noted in the table. Any disagreement was resolved by other reviewers also.

#### Data analysis

Meta-analysis was performed using RevMan 5.4 software, using a fixed-effect model. P-value < 0.05 was considered statistically significant. The effect of variables was measured in the Odds ratio between two groups for all outcomes in the included studies. Statistical heterogeneity across studies was assessed using the I^2^ test, and the values less than 40% were considered to be indicative of might not be significant heterogeneity.^[22]^ Subgroup analysis could be done for four ANC only if statistical heterogeneity is higher than 40%. The cluster randomized controlled trial’s design effect has been calculated to calculate the effective sample size and, accordingly, events in both treatment and control groups.^[23]^ Funnel plots have been created to show an effect estimate against its standard error for each outcome.

#### Patient and Public Involvement statement

This research does not have direct patient and public involvement. We used the relevant data from available literature only.

## RESULTS

A total of 319 articles were identified after searching through various databases. A reference list of selected articles was also explored to find the relevant articles. Further, two hundred eighty-seven articles were screened after removing duplicate articles. Thirty-two full-text articles were assessed for eligibility, and 26 articles were excluded due to different study designs, conducted in high-income countries, different outcomes, non-comparable control groups, and only protocol availability. Only six articles were included for the final meta-analysis. (Figure1 -PRISMA, Flow Chart)

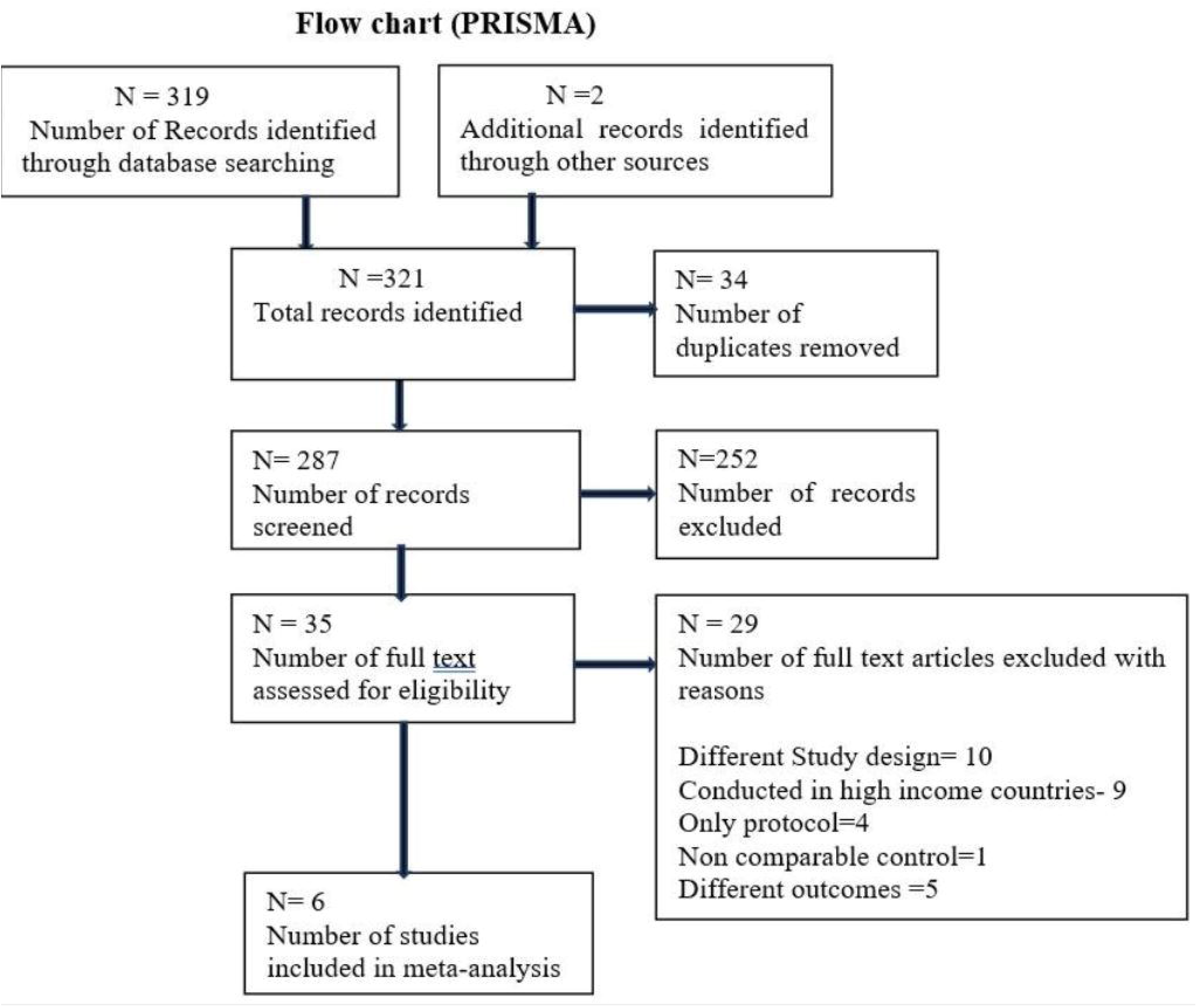

Total participants for the analysis of four or more ANC outcome (1115 interventional and 1133 control group), TT immunization outcome (601 interventional and 611 in the control group), compliance to iron therapy outcome (369 in interventional and 412 in the control group) and postnatal attendance outcome (1499 in interventional and 1340 in the control group) were included. The characteristics of the included studies were shown in table 1. Mudey, 2015^[12]^, Fedha, 2014^[13]^, Kebede, 2019^[15]^, Bangal, 2017^[16]^ and Adanikin, 2013^[17]^ included an intervention as SMS reminders and Lund, 2014^[14]^ included intervention as an automated short messaging service (SMS) system and a mobile phone Voucher system. All studies had comparators as standard care. Lund, 2014^[14]^ and Kebede, 2019^[15]^ were the cluster randomized controlled trials, and the sample size has been recalculated for the actual sample size considering the study’s design effect. We have performed sensitivity analysis and excluded studies due to methodological issues and different target populations. ^[24, 25]^ The summary of findings has been shown in table 2. This table has displayed the relative effect (95% CI) in the Odds ratio with events per 1000 participants in both arms. The certainty of evidence and grade have been shown as high, moderate, and low for each outcome variable. Forest plot [Figure -2, Figure- 3] shows the comparison between mHealth intervention and the control group with pooled data and favors mHealth intervention’s effectiveness in utilizing antenatal and postnatal care. Forest plot [Figure-2] favors the mHealth intervention group, (OR= 1.89 (95% CI- 1.49-2.19, I^2^= 33%) for the outcome of four or more antenatal attendance, forest plot favors the mHealth intervention group, (OR=1.63, 95% CI-1.17-2.27, I^2=^ 0%) for the outcome TT immunization of pregnant mothers, forest plot favors the mHealth intervention group, (OR=1.88, 95% CI- 1.18-3.00, I^2^=0%) for the outcome of compliance with iron supplementation and forest plot [Figure-3] favors the mHealth intervention group, odds ratio 2.54 (95% CI-2.15- 2.99, I^2^= 36%) for the outcome of postnatal attendance of delivered mothers. A funnel plot has been created to evaluate the publication bias and show an effect estimate against its standard error for antenatal care utilization outcomes. [Figure 4].

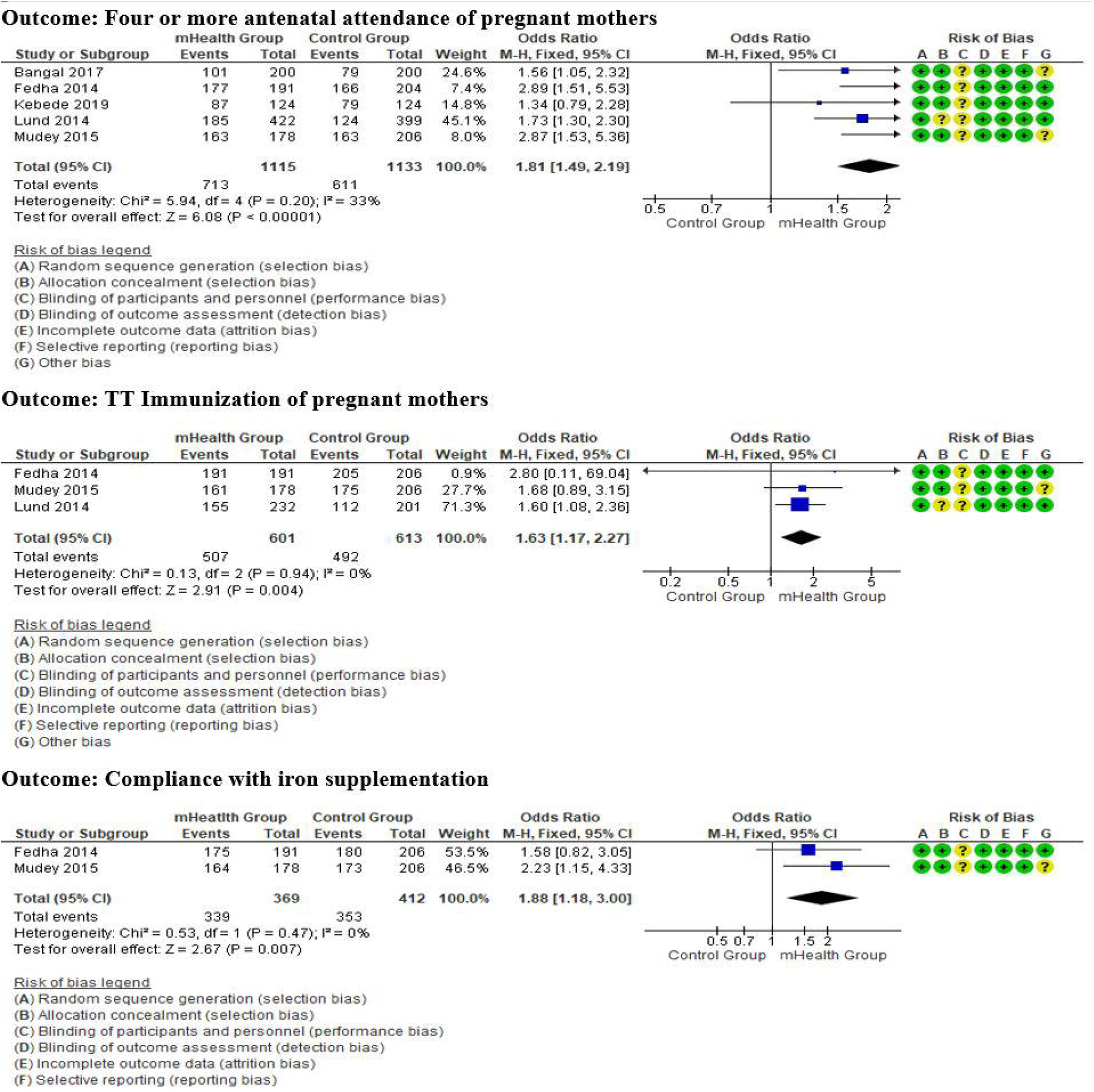

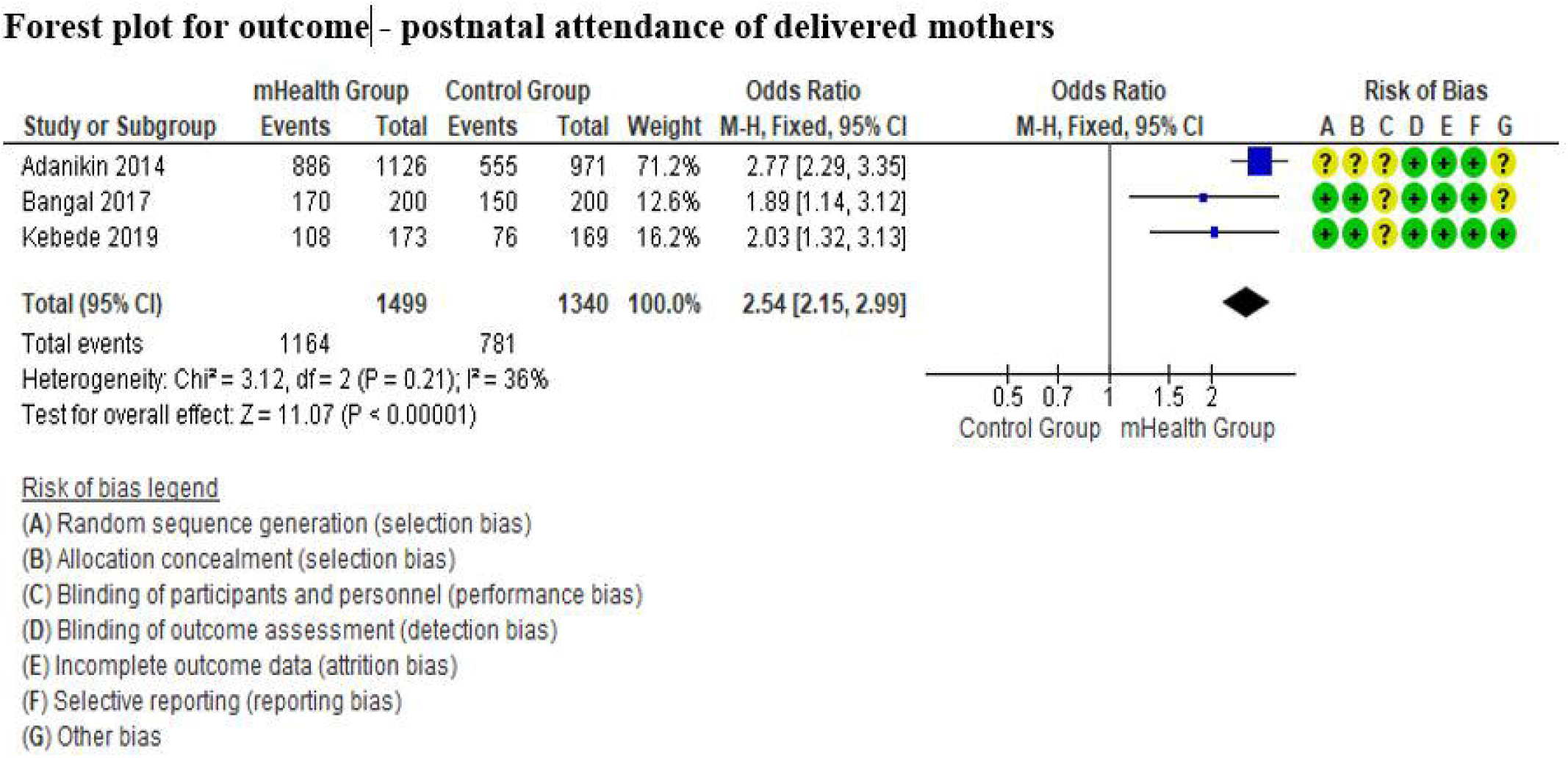

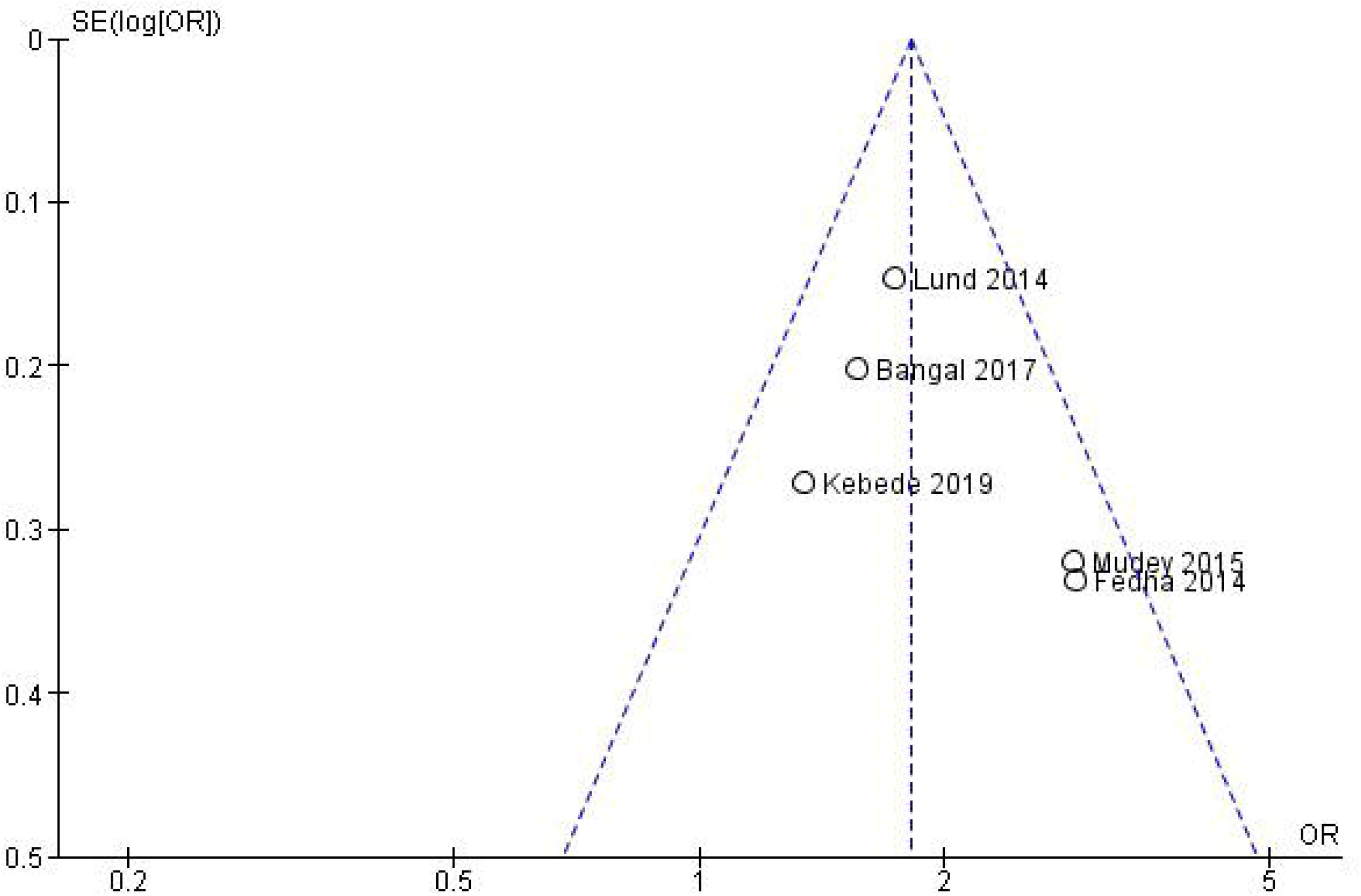

**Table 1:** Study characteristics.

| **Table 1: - Study characteristics** |  |  |  |  |  |
| --- | --- | --- | --- | --- | --- |
| **Study** | **Participants** | **Setting** | **Intervention** | **Comparator** | **Outcome** |
| Adanikin, 2013<sup>[17]</sup> | 1126 interventions and 971 controls | Nigeria | SMS reminders | standard care | postnatal care |
| Bangal, 2017<sup>[16]</sup> | 200 interventions and 200 controls | India | SMS reminders | standard care | Four or more ANC, TT immunization, Postnatal care |
| Fedha, 2014<sup>[13]</sup> | 191 interventions and 206 controls | Kenya | SMS reminders | standard care | Four or more ANC and compliance to iron therapy |
| Kebede, 2019<sup>[15]</sup> | 173 interventions and 169 controls | Ethiopia | SMS reminders | standard care | Four or more ANC, Postnatal care |
| Lund, 2014<sup>[14]</sup> | 1311 interventions and 1239 controls | Zanzibar | An automated short messaging service (SMS) system and a mobile phone voucher system | standard care | Four or more ANC and TT immunization |
| Mudey, 2015<sup>[12]</sup> | 178 interventions and 206 controls | India | SMS reminders | standard care | Four or more ANC and compliance to iron therapy |

**Table 2.** GRADEpro Summary of findings table

| Table:- 2 GRADEpro Summary of findings table |  |  |  |  |  |
| --- | --- | --- | --- | --- | --- |
| Outcomes | Anticipated absolute effects* (95% CI) |  | Relative effect (95% CI) | No of participants (studies) | Certainty of the evidence (GRADE) |
|  | Events with Control Group | Events with mHealth intervention |  |  |  |
| four or more Antenatal check up | 539 per 1,000 | **683 per 1,000**<br>(626 to 734) | **OR 1.84**<br>(1.43 to 2.36) | 2248<br>(5 RCTs) | ⊕⊕⊕○<br>MODERATE <sup>a</sup> |
| TT Immunization | 805 per 1,000 | **870 per 1,000**<br>(827 to 903) | **OR 1.62**<br>(1.16 to 2.26) | 1212<br>(3 RCTs) | ⊕⊕⊕○<br>MODERATE <sup>b</sup> |
| Compliance with iron supplementation | 857 per 1,000 | **918 per 1,000**<br>(876 to 947) | **OR 1.88**<br>(1.18 to 2.99) | 781<br>(2 RCTs) | ⊕⊕○○<br>LOW <sup>b, c</sup> |
| Postnatal care utilization | 583 per 1,000 | **770 per 1,000**<br>(722 to 811) | **OR 2.39**<br>(1.86 to 3.07) | 2839<br>(3 RCTs) | ⊕⊕○○<br>LOW <sup>b, c</sup> |

## DISCUSSION

Full antenatal care is a term used to deliver appropriate antenatal care to the pregnant mothers, inclusive of four or more antenatal check-ups, at least one tetanus toxoid injection with consumption of a minimum of 100 iron-folic acid tablets ^[2]^, which helps to promote the delivery of a healthy baby to a healthy mother. ^[3]^ Unfortunately, it is observed that a woman in low-income countries has 120 times higher risk of mortality due to pregnancy and childbirth-related causes compared to higher-income countries. ^[4, 6]^ Although studies have shown the effectiveness of mHealth intervention towards maternal and child health outcomes. ^[12-17]^ In the present meta-analysis, we have pooled the data of selected studies in low- and middle-income countries to create or strengthen the evidence for mHealth’s effectiveness towards antenatal and postnatal care utilization. This meta-analysis detected that m-health has the potential to increase antenatal and postnatal attendance compared to the standard approach, although the level of evidence was moderate. Results of this meta-analysis depicted the significant increase in four or more antenatal care attendance, TT immunization, compliance to iron supplementation, and postnatal care attendance among those pregnant mothers who received m-health intervention in comparison to those who did not receive such intervention. These findings were supported by two systematic reviews performed in low- and middle-income countries, although with low and moderate evidence levels. ^[26, 27]^ One systematic review suggests that mHealth intervention can be more effective in enhancing maternal and child health services if we target the mothers during antenatal and postnatal periods.^[27]^ If we incorporate the findings of a meta-analysis conducted in China, it also clarifies that mHealth apps with social media can also enhance maternal health.^[28]^ One of the evidence from the systematic review strengthens the findings that mHealth nutrition interventions also help in improving the dietary intake and nutritional status of pregnant mothers.^[29]^ However, a mixed-method study performed in South Africa shows that mHealth intervention was not useful in improving pregnant mothers’ health knowledge but useful in motivating their self-reported behavior to seek medical services. ^[30]^

In the rural population, a community-based randomized controlled trial in Ethiopia supports the positive contribution of SMS-based intervention towards maternal health. We even explored this study but excluded while sensitivity analysis due to methodological issues and target populations as community health workers and health extension workers. ^[24]^ A randomized controlled trial shows the higher satisfaction level of pregnant women who received SMS during their antenatal duration than the routine antenatal care group. This study also presented the lower anxiety level among pregnant mothers during their antenatal period, but they did not notice any difference in pregnancy outcomes.^[31]^ A controlled quasi-experimental study in Tanzania on pregnant women in improving pregnant mothers’ knowledge on danger signs and birth preparedness also found a significant difference between mHealth and standard care. ^[32]^ Ultimately it will one step towards improving antenatal and postnatal care attendance. As we have discussed, systematic reviews and meta-analyses have been conducted to examine mobile intervention’s effectiveness on different maternal and child health outcomes. However, our meta-analysis findings contribute to mHealth intervention’s effectiveness in utilizing full antenatal care and postnatal care among pregnant mothers.

### Strengths and limitations of this study

This meta-analysis creates an evidence for the effectiveness of mHealth with pooled data of interventional studies with limited sample sizes. This meta-analysis adheres with the Preferred Reporting Items for Systematic Review and Meta-Analysis for Protocols 2009 to ensure the quality of reporting the results. Sensitivity analysis identified possible reasons for heterogeneity among studies. As we included studies from LMICs, so results can be generalized for the respective population. mHealth as an intervention is a broad term that created heterogeneity also.

## Conclusion

This meta-analysis concluded that m-health intervention has the potential to increase the utilization of full antenatal and postnatal care compared to the standard approach, although the level of evidence was moderate. Technology is changing, but even with limited support like SMS, there was an improvement in antenatal and postnatal service utilization. It might have a possible solution to enhance maternal and child health services to pregnant mothers and reduce maternal and neonatal mortality. Further studies are required with experimental trials or cluster randomized controlled trials to assess the feasibility and cost-effectiveness of mHealth intervention in community settings with the next step by involving the government health system to implement these findings.

## Data Availability

Data included in the meta-analysis have been presented in the manuscript. Additional information related to the study has been uploaded as supplementary files.

## Contributions details

Study conception/design: Yadav P, Kant R,

Data collection: Yadav P, Kant R, Kishore S, Barnwal S

Data analysis, and interpretation: Yadav P, Kumar R, Khapre M, Barnwal S

Drafting Manuscript: Yadav P

Revising manuscript: Yadav P, Kant R, Kishore S, Kuamr R, Khapre M, Barnwal S

Approval of final version of the manuscript for publication: Yadav P, Kant R, Kishore S, Khapre M, Kuamr R, Barnwal S

Responsibility for accuracy and integrity of all aspects of research involvement in drafting or revising the manuscript: Yadav P, Kant R, Kishore S, Khapre M, Kuamr R, Barnwal S

## Patient consent for publication

**Not Applicable**

## PRISMA Checklist

Supplementary file 1

## The original protocol for the study

Supplementary file 2

## Data availability statement

Data included in meta-analysis have been presented in the manuscript. Additional information related to the study has been uploaded as supplementary files.

